# Strategies and action points to ensure equitable uptake of COVID-19 vaccinations: A national qualitative interview study to explore the views of undocumented migrants, asylum seekers, and refugees

**DOI:** 10.1101/2021.04.12.21255313

**Authors:** Anna Deal, Sally E Hayward, Mashal Huda, Felicity Knights, Alison F Crawshaw, Jessica Carter, Osama B Hassan, Yasmin Farah, Yusuf Ciftci, May Rowland-Pomp, Kieran Rustage, Lucy Goldsmith, Monika Hartmann, Sandra Mounier-Jack, Rachel Burns, Anna Miller, Fatima Wurie, Ines Campos-Matos, Azeem Majeed, Sally Hargreaves, on behalf of the ESCMID Study Group for Infections in Travellers and Migrants (ESGITM)

## Abstract

**Introduction:** Early evidence confirms lower COVID-19 vaccine uptake in established ethnic minority populations, yet there has been little focus on understanding vaccine hesitancy and barriers to vaccination in migrants. Growing populations of precarious migrants (including undocumented migrants, asylum seekers and refugees) in the UK and Europe are considered to be under-immunised groups and may be excluded from health systems, yet little is known about their views on COVID-19 vaccines specifically, which are essential to identify key solutions and action points to strengthen vaccine roll-out.

**Methods:** We did an in-depth semi-structured qualitative interview study of recently arrived migrants (foreign-born, >18 years old; <10 years in the UK) to the UK with precarious immigration status between September 2020 and March 2021, seeking their input into strategies to strengthen COVID-19 vaccine delivery and uptake. We used the ‘Three Cs’ model (confidence, complacency and convenience) to explore COVID-19 vaccine hesitancy, barriers and access. Data were analysed using a thematic framework approach. Data collection continued until data saturation was reached, and no novel concepts were arising. The study was approved by the University of London ethics committee (REC 2020.00630).

**Results:** We approached 20 migrant support groups nationwide, recruiting 32 migrants (mean age 37.1 years; 21 [66%] female; mean time in the UK 5.6 years [SD 3.7 years]), including refugees (n = 3), asylum seekers (n = 19), undocumented migrants (n = 8) and migrants with limited leave to remain (n = 2) from 15 different countries (5 WHO regions). 23 (72%) of 32 migrants reported being hesitant about accepting a COVID-19 vaccine and communicated concerns over vaccine content, side-effects, lack of accessible information in an appropriate language, lack of trust in the health system and low perceived need. Participants reported a range of barriers to accessing the COVID-19 vaccine and expressed concerns that their communities would be excluded from or de-prioritised in the roll-out. Undocumented migrants described fears over being charged and facing immigration checks if they present for a vaccine. All participants (n = 10) interviewed after recent government announcements that COVID-19 vaccines can be accessed without facing immigration checks remained unaware of this. Participants stated that convenience of access would be a key factor in their decision around whether to accept a vaccine and proposed alternative access points to primary care services (for example, walk-in centres in trusted places such as foodbanks, community centres and charities), alongside promoting registration with primary care for all, and working closely with communities to produce accessible information on COVID-19 vaccination.

**Conclusions:** Precarious migrants may be hesitant about accepting a COVID-19 vaccine and face multiple and unique barriers to access, requiring simple but innovative solutions to ensure equitable access and uptake. Vaccine hesitancy and low awareness around entitlement and relevant access points could be easily addressed with clear, accessible, and tailored information campaigns, co-produced and delivered by trusted sources within marginalised migrant communities. These findings have immediate relevance to the COVID-19 vaccination initiatives in the UK and in other European and high-income countries with diverse migrant populations.

**Funding:** NIHR

## Introduction

Ethnic minority and migrant populations have been disproportionately affected by the COVID-19 pandemic in the UK (1-3), yet early evidence suggests low intent and uptake of the COVID-19 vaccination (4-9). Specifically, numerous UK surveys show low vaccine intent in Black, black British and Asian ethnic minorities in the UK. One recent survey showed only 57% people from Black, Asian, and minority ethnic backgrounds would accept a COVID-19 vaccine, compared to 79% of White respondents (10). Recently published UK vaccine uptake data for the over 70’s age-group has shown lower vaccination rates in ethnic minority groups, particularly Black African (58.8%) compared to White British 91.3% (11). Lack of trust in government or health systems, social exclusion, and long running issues of discrimination have been highlighted as contributors to COVID-19 vaccine hesitancy in minority ethnic groups (4-6, 9). None of the currently available datasets, however, give insight specifically into the views of migrants (defined as foreign-born) (12). In particular, recently arrived migrants with precarious immigration status such as undocumented migrants, asylum seekers and refugees, who are known to face many unique barriers and even exclusion from health systems, have not been well considered in research to date around the COVID-19 vaccine. Many are concerned that vulnerable groups, including precarious migrants, homeless populations, and Roma communities, as well those living in highly deprived areas, may not be reached in the COVID-19 vaccine roll-out without specific interventions to facilitate engagement with these communities in order to strengthen delivery and uptake (13, 14).

In the UK, the COVID-19 vaccination roll-out will be predominantly carried out through existing healthcare services, however, large numbers of migrants are currently excluded from these for a number of reasons. These include perceived or true lack of entitlement to access healthcare, fear around charging or links to immigration services and poor understanding of the system, often compounded by language barriers (15, 16). These barriers have been exacerbated in many cases by increasing digitalisation of healthcare services during the pandemic (17, 18). Adult migrants are widely excluded from vaccination services on arrival to the UK and Europe, due to structural and policy shortfalls in engaging them in catch-up vaccination campaigns (19, 20), even though ensuring high levels of coverage and equitable access are key priorities of the European Vaccine Action Plan and the Sustainable Development Goals (SDGs) (21). Specifically, undocumented migrants or those with limited leave to remain, asylum seekers and refugees, those residing in temporary asylum accommodation, detention centres, and other high-risk settings, as well as several groups of low-skilled labour migrants, are also often highly marginalised and excluded from health and vaccination systems, yet their views are rarely sought to inform policy and practice. Around 1.2 million undocumented migrants alone may be currently residing in the UK (22), of whom many may not be registered with primary healthcare services where COVID-19 vaccination is currently being delivered. Policies aiming to restrict access to healthcare for overseas visitors in the UK, as part of the political ‘hostile environment’ towards undocumented migrants, which has included patient data sharing agreements between the health service and the Home Office for immigration enforcement purposes (23), have caused a lack of trust and confusion around entitlement to healthcare among both NHS staff and patients (24-26). This has resulted in precarious migrants only accessing services when in urgent need and avoiding preventative health services such as vaccination (16), with calls for immigration data sharing and immigration checks at health services to be suspended during the pandemic. Increased social exclusion during the pandemic may have exacerbated long-running issues of mistrust and mutual lack of understanding between public health services and migrants, impacting on their willingness to present to health services to get vaccinated (18, 27). Precarious migrants have been reported to be avoiding hospitals for fear of charging if they are negative for COVID-19 (28), despite Public Health England specifically stating that COVID-19 vaccines are free of charge and no immigration checks will be carried out (29).

For COVID-19 vaccination strategies to be effective in the UK, the vast majority of the adult population will undoubtedly need to be vaccinated now and in the future (30), including migrants and other marginalised groups who may have a range of risk factors related to COVID-19. It is essential that we better understand factors affecting vaccine intention and acceptance among precarious migrant groups to better understand their views or concerns and to define strategies to ensure equitable access and delivery. We therefore did an in-depth qualitative interview study of recently arrived (<10 years) precarious migrants to explore views on the COVID-19 vaccine, including barriers to access, seeking their input into defining action points and developing solutions to strengthen delivery and uptake in marginalised migrant communities.

## Methods

### Study and Interview Design

We used a qualitative methodology consisting of in-depth semi-structured interviews, to explore the perspectives of recently arrived migrants (residing in the UK <10 years). We specifically aimed to recruit migrants with precarious immigration status, including refugees, asylum seekers and undocumented migrants (including visa overstayers, refused asylum seekers, and others lacking documentation), and individuals with limited leave to remain (migrants on temporary visas, with no recourse to public funds). The research design was ideally suited to the exploratory nature of the research questions, seeking to reveal perspectives and understanding of the participants in this novel area of investigation. Interviews were carried out between 4^th^ September 2020 and 8^th^ March 2021. Topic guides were developed by the research team comprising AD, SH, SMJ, AC, SEH (academic researchers) and JC, FK (General Practitioners), with input from the Migrant Health Research Groups’ Project Board, including migrant representatives from a range of countries, ages and backgrounds. The topic guide was developed through iterative cycles and informed by the situation of the pandemic and the progression of the UK COVID-19 vaccine roll-out and piloted prior to the study starting. Participants were asked broadly about their experiences of the pandemic and their views concerning the COVID-19 vaccine, including barriers and facilitators to access, as well as a direct line of questioning pertaining to whether they would accept a COVID-19 vaccine or not. In addition, we sought their views around delivery strategies for their communities and solutions to ensure equitable uptake.

### Participant Recruitment

Recently arrived migrants were recruited using purposive and snowball sampling, with the aim of recruiting participants from a broad range of nationalities, migrant statuses and age groups. Adverts for the study and participant information sheets were circulated to 20 UK-based migrant support groups and on social media. Those who expressed an interest in taking part were contacted by telephone and the study was explained to them with interpreters available on request.

### Ethics and informed consent

Ethics was granted by St George’s, University of London Research Ethics Committee (REC 2020.0058 and 2020.00630). Participant information sheets were circulated, and informed consent was acquired in writing prior to arranging a telephone interview.

### Data Collection and Analysis

In-depth semi-structured interviews were conducted by telephone (by AD, SEH) and lasted 30-90 minutes. Participants were compensated with an online shopping voucher (worth £37), as per INVOLVE NIHR criteria for participant involvement in research studies (31). Interviews were audio-recorded then transcribed verbatim; transcripts were checked for accuracy and anonymised. Data collection ended when data saturation was reached, and no novel concepts were arising (13). Data collection and theme development took place concurrently and continued until the team agreed unanimously that saturation, at a thematic level had been reached. Data were then analysed using the thematic framework technique (32) in NVIVO 12. We used the ‘Three Cs’ model of vaccine hesitancy, which focuses on issues pertaining to confidence in the vaccine, complacency, and convenience which are considered to influence an individuals’ views on whether to have a vaccine or not (33, 34). Finally, we did a sub-analysis exploring views and levels of hesitancy among migrants interviewed before (September and November 2020) and after (between January and March 2021) the beginning of the COVID-19 vaccination roll-out in the UK.

## Results

### Participant demographics

We carried out 32 interviews (4^th^ September 2020 to 8^th^ March 2021), with 17 interviews done between September and November 2020 and 15 between January and March 2021. Participants reported their migration status as seeking asylum (n = 19), refugees (n = 3), undocumented (n = 8) and limited leave to remain (n = 2). The mean age across the study sample was 37.1 years (SD: 7.6 years); 21 (66%) participants were female. The mean duration of stay in the UK was 5.6 years (SD 3.1 years), with 17 (53%) participants who had resided for five or less years in the UK, and 15 (47%) who had stayed been in the UK 5-10 years. Multiple nationalities were represented among respondents, with participants coming from five WHO regions and 15 different countries. Participant demographics are further described in Table 1.

**Table 1.** Demographics of study participants (n = 32)

| Characteristic | n (%) |
| --- | --- |
| **Migrant status** |  |
| Asylum seekers | 19 (59%) |
| Refugees | 3 (9%) |
| Undocumented | 8 (25%) |
| Limited leave to remain | 2 (6%) |
| **Age (years), mean (SD)** | **37.1 (7.6)** |
| < 35 | 11 (34%) |
| 35-50 | 19 (59%) |
| >50 | 2 (6%) |
| **Gender** |  |
| Female | 21 (66%) |
| **WHO region of Origin** |  |
| African (Mauritius, Nigeria, Uganda, Zimbabwe, Other unstated) | 11 (34%) |
| The Americas (Venezuela) | 1 (3%) |
| Eastern Mediterranean (Afghanistan, Egypt, Iraq, Pakistan, Palestine) | 11 (34%) |
| European (Albania, Kyrgyzstan, Turkey, Ukraine, Other) | 5 (15%) |
| South East Asian (Sri Lanka) | 4 (12%) |
| **Time since arrival in the UK (years), mean (SD)** | **5.6 (3.1)** |
| <2 | 3 (9%) |
| 2-5 | 14 (45%) |
| 5-10 | 15 (47%) |
| **Location in the UK** |  |
| England - London | 14 (44%) |
| England - North East | 7 (22%) |
| England - Other/Unknown | 10 (31%) |
| Wales | 1 (3%) |

### General views on barriers to COVID-19 vaccination access for precarious migrants

#### Lack of defined access points

Participants raised similar concerns around how they were going to access the COVID-19 vaccine, stemming from concerns around existing access issues to primary care, such as language barriers, trust issues, or perceived lack of entitlement. However, some barriers identified varied by migrant status, with undocumented participants and those with limited leave to remain often reporting different barriers to refugees and asylum seekers. Concerns were raised that some precarious migrants, particularly those who are undocumented, are not registered at a GP practice and will therefore be excluded from the COVID-19 vaccine roll-out.

*“Some of the asylum-seekers and the refugees, they don’t have a GP, so I don’t know how the government will help out with that. If the government can speak with the charities, because a lot of these refugees and asylum-seekers, they use different charities” -Asylum seeker 16*

Participants stated that they have historically relied on charities and walk-in centres for healthcare and help with GP registration, and that the discontinuation or digitalisation of many of these services during lockdown has heavily affected them, impacting on their ability to register with a GP. Very recently arrived migrants (<2 years in the UK) described difficulties registering with a GP using the digital NHS system in place during the pandemic. Additional barriers to access through primary care are further discussed in the vaccine hesitancy “convenience” section below.

#### Distrust of health and vaccination systems

Many participants described a lack of trust in both healthcare or wider governance systems, with bad previous experiences and anecdotes from friends or family often a contributing factor. In particular, difficulty understanding the NHS system on arrival and poor treatment by staff during registration processes was reported as a factor affecting trust in healthcare services for asylum seekers and refugees, which may impact on COVID-19 vaccine uptake in these groups. Experiences of being charged for healthcare, particularly maternity services, amongst undocumented migrants has led to a lack of trust in government messaging and perpetuated fear around charging and immigration checks, which they felt could have implications for vaccine roll-out. No migrants interviewed after the recent government announcement (n = 10, 8^th^ February – 8^th^ March 2021), stating that COVID-19 vaccinations will be given without immigration checks taking place, reported being aware of this.

*“So, when I went to the GP where I still use now to register, the woman there, the lady there was so harsh on me. She wouldn’t… It was just like I’m a trash of rubbish in her face. She started demanding that I should bring this, I should bring that” - Asylum seeker 19*

*“They should campaign for the free giving of the vaccine without payment. And government should not come and hunt you that you are owing them. Because you took the vaccine you are owing them” -Undocumented 6*

*“They don’t need to put the word documents [in Covid vaccine adverts] because*… *what if I don’t have it, I’m undocumented. And you said okay come on have your vaccine, we’re not going to check you*… *I won’t go because I don’t know to what extent is true. It might be a ploy to get people to come” - Asylum seeker 17*

#### Feeling abandoned during the pandemic

Many participants described feeling abandoned by the government and health services during the pandemic, leading to concerns that they would also be ignored or excluded from the vaccine roll-out. Participants reported substantial strain on their mental and physical health as well as financial and social issues. Those seeking asylum often reported major delays in their asylum applications and being left in temporary shared accommodation, where social distancing was not possible, perpetuating their sense of being abandoned.

*“I listened that it [COVID-19 vaccinations] will come in UK. They will give the British people first and we asylum seekers will come in the end” Asylum seeker 15*

*“It boils down to the neglect in human race like neglecting, what would I call it, vulnerable people, people with low standard of life that you are not taking serious… When you’re saying don’t touch this, and don’t touch this, don’t share, people are still in shared accommodation. They never care for the less privileged” - Asylum seeker 19*

*“I would say mainly we wouldn’t trust it [a COVID-19 vaccine], just because they [the government] don’t care” -Refugee 2*

### Views on COVID-19 vaccination: Confidence, Complacency and Convenience

Participants reported a range of views on the COVID-19 vaccines, ranging from complete acceptance to fear and distrust, which are summarised in Figure 1. When we explored differences between pre-(September – November 2020) and post-announcement (January – March 2021) of vaccination deployment, we found participants interviewed earlier in the pandemic were generally more concerned about the vaccine and influenced by misinformation than those interviewed after the start of the vaccination roll-out. Those who had a more positive view of the COVID-19 vaccine (would accept a vaccine or were unsure but leaning towards accepting, n = 17) often described seeing it as way of getting out of difficult living situations that the pandemic has imposed upon them. Twenty-three (72%) of 32 respondents were hesitant about accepting a COVID-19 vaccine, however, 10 (43.5%) among these reported leaning towards accepting.

**Figure 1.**
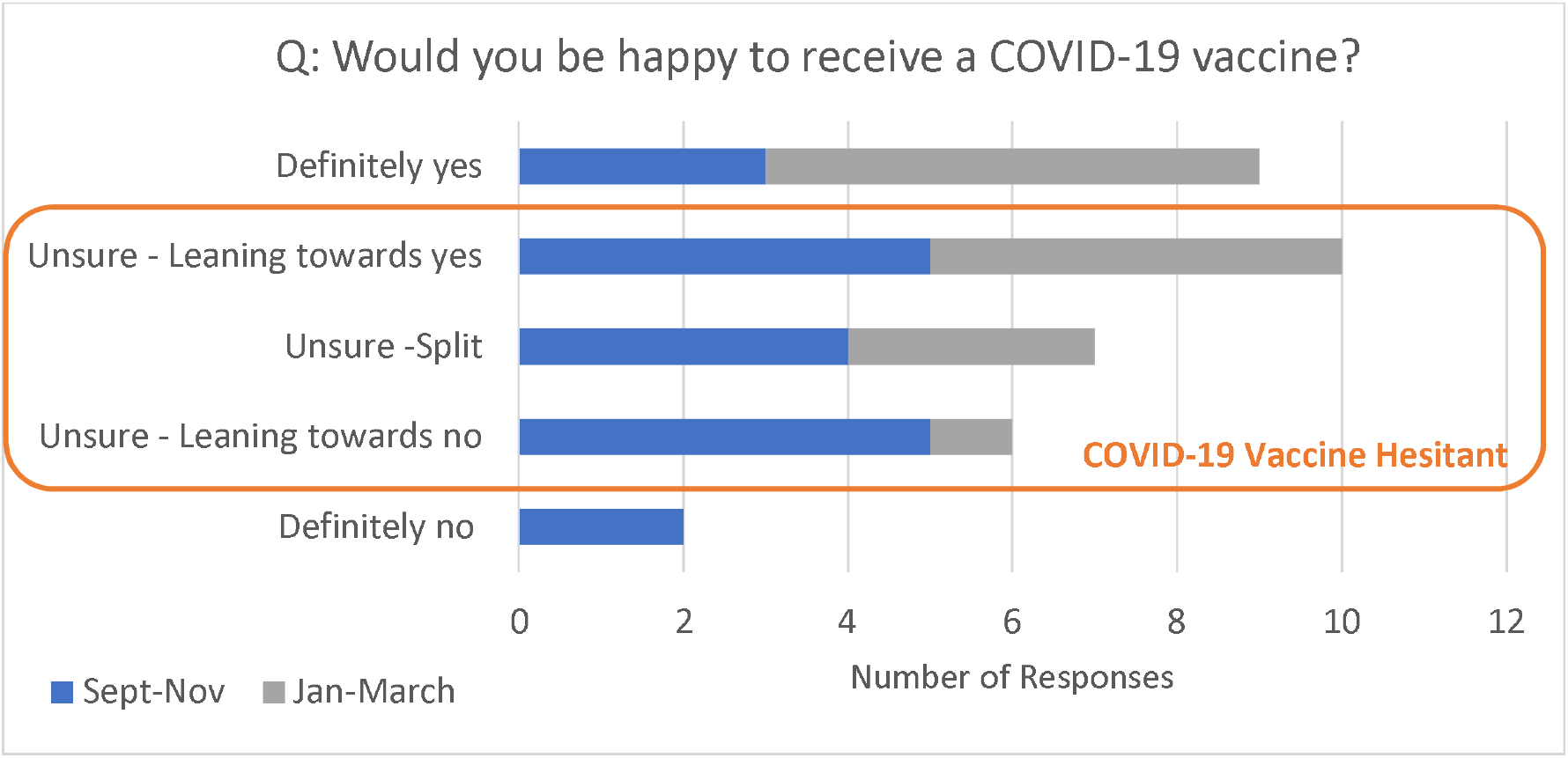
Participant responses (n = 32) on acceptance of a COVID-19 vaccine, categorised by time of interview.

#### Confidence in COVID-19 vaccination

Confidence was a key factor for the majority of those who stated they were, at the time of interview, hesitant about accepting a vaccine (n = 23). These included worries around potential side-effects and insufficient testing of the vaccine during clinical trials to ensure its safety. Some participants also described fears around theories based on misinformation, often originating from social media or word of mouth, with many describing feeling conflicted about which information sources to trust. Of those who stated they would definitely not accept a vaccine (n = 2), one said this was due to lack of clinical trials data and the other stated religious reasons (considered vaccines anti-Islamic). In general, those who described stronger feelings of social exclusion during the pandemic were more likely to express distrust in a vaccine.

*“For me, I would like to take the vaccine if that will make everything better. But the fake news is scaring me, so I don’t know. That is a problem. I don’t know if it’s real, I don’t know if it’s fake. When you take it, it will change the DNA… it will stop the person not having kids in future. A lot of stories are flying” - Asylum seeker 16*

*“It depends because you hear fake news everywhere… The only thing that one I heard about, and I still don’t know whether it’s fake news or not, but the only thing that worried me, like if they would relate the microchip with the vaccine” - Asylum seeker 2*

*“For me anything that’s going to ease the pandemic right now is very okay. If it’s really going to be effective, then why not? I’m just scared about it. I don’t know if it’s going to have a side effect. That bit is what I’m worried about” -Undocumented 7*

#### Complacency in COVID-19 vaccination

Several participants displayed complacency towards a COVID-19 vaccine, considering it to not be needed, and preferred to rely on either natural remedies, their immune system, or self-isolation to prevent them getting COVID-19 rather than having a vaccine. Views around complacency were often related to a lower trust in preventative medicine in general.

“*I don’t think that COVID will remain a threat for us for a long time*… *So why should I put extra artificial materials in my body? Then tomorrow something else will definitely come and I cannot be vaccinating myself against everything*… *I put something into my mouth only when it becomes too hurtful. I always delay it to the last moment. Like a high temperature or whatever pain, I try to cope myself… So, we use all the sources of our bodies first before taking something from outside” - Asylum seeker 7*

#### Convenience and COVID-19 vaccination

Participants stated that convenience would be a key factor in their decision on whether to accept a vaccine or not. The most commonly mentioned concern was ease of access, including having enough understandable information on where and when they would have to present as well as a preference for familiar settings requiring minimal travel. Those who reported having positive past experiences with their GP, particularly refugees and asylum seekers, mostly stated they would feel most comfortable receiving the vaccination in primary care. In contrast, those with undocumented status generally put more importance on being assured anonymity when presenting for the vaccination, and many preferred to access it through walk-in centres or trusted charities. Costs associated with the vaccine, both direct and indirect (e.g. travel), were also a major factor for many participants, with many unsure if the vaccine would be free despite existing government messaging that the vaccines will not be charged for. Participants stated that if they were confident there would be no associated costs, this would enable many more people within their communities to present for a vaccine.

*“People don’t have money now because of this what is going on. So, if you want to take it, government should make available for them. They should just make it equal for everybody to take the vaccine freely. Because some cannot afford to pay to take the vaccine” -Undocumented 6*

*“If you make it easier for everyone in this country whether registered in GP or you don’t have document, just come and maybe go to centre or go to GP… If they do it like that, most people will still go [to get their vaccine]” -Undocumented 5*

“*It depends on the distance. Because as I said, we are not able to offer ourself to transport, we cannot pay for it. At some points like some people they have health issue where they cannot walk so long. I mean long distance. So, it depends on the distance and in some conditions maybe it would be in the GP, in some others it would be the [walk-in vaccination] centre” -Undocumented 2*

#### Lack of information influencing views on COVID-19 vaccination

Many participants felt they had not had access to sufficient understandable information on the pandemic or on COVID-19 vaccines, with language barriers often brought up as an issue. In general, those feeling most abandoned or scared due to a lack of understandable, clear official information in the early stages of the pandemic were more likely to rely on word-of-mouth or social media (WhatsApp groups, Facebook) for subsequent information on the pandemic and the ongoing vaccination programme.

*“I don’t know how the government receive it [COVID-19 information] at first, because I think… like I said, because they are giving a lot of confusion to the society. They are passing a lot of confusion” - Asylum seeker 19*

*“What the government or NHS can do to improve, or to facilitate, or to help this lack of knowledge [around COVID-19 vaccines] is to communicate. Communication is very, very, very essential because lack of communication can just lead to disaster” - Asylum seeker 11*

Many participants who were currently hesitant about accepting a COVID-19 vaccine stated they would need more information before making their decision, preferably in their own language, on potential side effects of the vaccines, vaccine contents, summaries of clinical trial data, and when and how they would be invited for a vaccine. Several participants stated that they would like circulating misinformation to be directly addressed by official information sources (including the Government and the NHS). Many undocumented participants also described that they would need more information about whether documentation would be asked for at their access points (e.g. walk-in clinics), and felt that they currently lacked information in this area. A wide range of formats were suggested for improving information accessibility and reach, which are further explored in Table 2, including both traditional methods (leaflets, TV news channels, internet-based sources, posters) and more novel ideas (social media, community champions, existing charity networks). It was felt that a flexible, holistic approach, with information available in as many formats and languages as possible would be the most effective.

**Table 2.** A summary of key issues and proposed strategies for increasing vaccine access and uptake.

| Factors that may influence COVID-19 vaccine uptake | Strategies identified by participants to increase uptake | Quotes/Evidence |
| --- | --- | --- |
| Lack of trust in the health and vaccination system | • Use trusted groups or sources (NGOs, community groups) for communication<br>• Campaigns to increase trust in the primary care systems, without immigration checks or data sharing<br>• Facilitate access to primary care system, specifically registration with GPs<br>• Increased funding for and | *"the only way for the government to try and tackle that is to actually put a promise, a government promise to say that we promise that anyone can access it. And no information will be shared with the home office" -Undocumented 4*<br>*"If you have managed to accommodate the person and get the person to register to a GP, then if you ask him to do it [take the vaccine], he'll know that it's for my good. They are going to be happy that they are treating me now like other people, a priority. He will not feel abandoned* |
|  | collaboration with charities and community groups who act as a major source of healthcare and information to precarious migrants<br>• Ensure communications do not label specific groups as 'infectious' or 'hesitant', to avoid increasing stigmatisation and distrust | *anymore" -Undocumented 8*<br>*"Contact the local asylum organisations and groups so that staff can learn more about the vaccinations, about Covid things, and ask the asylum seekers their preference of communication and access and provision... Because it's difficult to communicate with somebody who's not understanding what you say" - Asylum seeker 11* |
| Lack of access points for the COVID-19 vaccine for migrants facing barriers to primary care - "Convenience" factors | • Use walk-in centres in safe and trusted places (such as community centres, charities, food banks, pharmacies, homeless services) for COVID-19 vaccine roll-out<br>• Campaigns to educate primary care staff on the right for migrants to access healthcare without documents | *"They should create a centre, like during elections... maybe they create in the street? Maybe they might have one or two or three centres there and people just walk in then instead of going to the GP or anything" - Asylum seeker 17*<br>*"Some of the asylum-seekers and the refugees, they don't have a GP, so I don't know how the government will help out with that. If the government can speak with the charities, because a lot of these refugees and asylum-seekers, they use different charities" - Asylum seeker 16*<br>*"The people that are not legal in the UK, they found that really hard to access any medical unless they registered in a homeless centre. And then they get the medical that they need through a homeless service unit" -Undocumented migrant 4*<br>*"They also have to meet the needs of the population with a wide range of literacy skills...* |
|  |  | *and provide ongoing training to NHS staff, on how to behave appropriately with patients who do not speak English” - Asylum seeker 11* |
| Low confidence in COVID-19 vaccines | • Accessible information required on side-effects, contraindications, contents, and to counter misinformation, see information section below<br>• Encourage the spread of information via word-of-mouth/social media by individuals who have taken the vaccine<br>• Specific campaigns to counter common misinformation circulating on the COVID-19 vaccine in communities, including healthcare workers, who may be more susceptible to misinformation due to lack of accessible information | *“I really want someone to come out and just say, this is it or this is what the vaccine is all about... explain more in details that this is what the vaccine contains, whatever you're hearing is false. This is it. The vaccine does not do this, it does not do that. They just clear everything so that people will be sure of what they're doing” - Asylum seeker 16*<br>*“I’ve been having concerns about this vaccine for Covid because of what I’ve been hearing from people talking... but if you are aware, you now know, and you have seen other people doing it, and then you see, okay, it’s preventing, it’s working, it’s been proven and they’re saying it on the TV and then I’ll go for it” - Asylum seeker 11*<br>*“Even today I want to call them because most of my friends said they have taken [the vaccine]. So, I want to take as well” - Asylum seeker 19* |
| Lack of accessible information sources around COVID-19 vaccine | • Holistic information campaigns in multiple formats and languages to increase reach<br>• Information should be tailored for specific groups, and presented in a sympathetic, culturally appropriate and | *“...we are in need of community champions because I’m worried, extremely worried, as people from difficult communities... They're not going to have the knowledge of who to contact and they will feel more scared [about the vaccine]” -Undocumented migrant 4*<br>*“You’ve also got your possibility of all the posters being put in a different language in the [asylum]* |
|  | understandable manner<br>Existing, effective and trusted channels should be used, for example charities, food banks, asylum hotels, TV channels, GP practices.<br>Local community champions should act as an information point for both those in their community and by those designing tailored information campaigns, with sufficient training to ensure effective delivery of methods. | *hostel, which will basically be very effective. They have posters for everything in there in all the languages” - Asylum seeker 14*<br>*“I think it would be useful to have connection with the local communities, like either community leaders, or the places that people access... religious places or something like that, to make it easier to have a connection with them, and make them aware of the vaccines” -Refugee 2*<br>*“Especially I am not reading letters. But I don’t know, other people, what they do. But I don’t read. I say, oh my God, what is this? Too much English, just put it there” - Asylum seeker 18* |

*“They should say is it going to have a side effect. This is what the outcome and everything… The only thing I know so far is from my GP actually, that there is a priority and they’re giving from the older age” -Undocumented migrant 7*

“*The prime minister has said we want everybody to be vaccinated. But he did not say no information will be given to the home office. He did not give that safety blanket and security to people that are afraid to give information away. So, this is my fear” -Undocumented migrant 4*

*“Some people can speak and some they can’t understand about the English… I suggest that all these things should be in other languages as well” -Undocumented migrant 2*

### Strategies to facilitate equitable COVID-19 vaccine uptake in precarious migrants

Participants described a wide range of strategies that they considered would be useful to either encourage them to accept a vaccine for themselves, or to increase the accessibility and reach of the vaccination programme for other precarious migrants in their communities. Participants highlighted strategies to combat the four main issues presented as hindering COVID-19 vaccine access for precarious migrants, namely, trust in the system, lack of defined vaccine access points for those with restricted access to healthcare, low vaccine confidence, and lack of accessible information on COVID-19 vaccines, migrants’ entitlement to access a vaccine, and if it would be free. The proposed strategies are described in detail in Table 2, and include using trusted groups or sources (NGOs, community groups) for communication, and to use these same groups as access points for COVID-19 vaccine delivery (for example, as hosts for walk-in centres). In addition, participants suggested campaigns to increase awareness of entitlement to primary care were also required. Increased funding for, and collaboration with, charities and community groups who act as a major source of healthcare and information to precarious migrants was also highlighted.

## Discussion

This is the first attempt to explore the views of a diverse range of precarious migrants on COVID-19 vaccine hesitancy and barriers and facilitators to vaccine uptake. Most participants (23 [72%] of 32) reported feeling hesitant about accepting a COVID-19 vaccine. Reasons given included concerns over vaccine content, side effects, lack of information or low perceived need, suggesting hesitancy could be easily addressed with clear, accessible and tailored information campaigns. Concerns were expressed that migrants may be excluded from the vaccine roll-out, and that migrants not registered with the health system had no access point for the vaccine. In addition, a wide variety of barriers to the vaccine were highlighted, including lack of registration with primary care services, fears over charging for the vaccine, data sharing between the health service and immigration enforcement, lack of information on alternative access points, and other issues relating to convenience. Our data suggest that the campaign allowing undocumented migrants to get a COVID-19 vaccine without immigration checks and free of charge may need to be more effectively communicated. A range of strategies and solutions were proposed by respondents to increase vaccine uptake, including culturally and linguistically accessible information campaigns in a range of formats and languages, innovative, trusted and well-defined access points, more flexible entry points to primary care, and increased collaboration with charities or groups already working with affected communities. These findings have direct implications for policy and practice during the current roll-out in the UK, but also will be salient for other routine vaccination campaigns, as migrants are known to be an under-immunised group generally (35). Our findings also highlight the consequences of excluding vulnerable groups from health systems and re-emphasize the importance of universal access to healthcare and effectively engaging with communities when formulating policy responses, which become particularly pertinent in the context of public health emergencies.

We found that precarious migrants may face a broad range of barriers to COVID-19 vaccination access, particularly those with undocumented status and others who are not registered with primary healthcare services. Lack of trust in authorities was a key theme, as well as concerns around immigration checks or other unwanted questions from healthcare providers if they present for a COVID-19 vaccine. These concerns were often based on previous experiences of charging by the NHS, poor treatment by NHS staff and the current hostile political environment that has embedded immigration enforcement within public services such as the health system (through mechanisms such as data sharing), which have been previously well-documented as barriers in access to healthcare (16, 24). The UK government announced in early February that undocumented migrants can register with a GP to get a COVID-19 vaccine without facing immigration checks (36), however, no statements have been made around whether this may lead to data sharing or immigration enforcement in the future. Furthermore, none of the participants interviewed in this study post-announcement (n = 10; 8^th^ February - 8^th^ March 2021) were aware of this when directly asked, and several were unsure if the vaccines would be free, despite previous statements to this effect (29). This suggests that current pathways used to disseminate such information are not effective in reaching their target audience, potentially due to language barriers. Doctors of the World have recently produced information sheets for migrants on the COVID-19 vaccine in 30 languages (37), however, only 6% of Governments in Council of Europe countries produced information on healthcare entitlement during the pandemic in common migrant languages, limiting information reach (38). There have been several recent initiatives to support marginalised groups access the COVID-19 vaccine, including mobile COVID-19 vaccination services targeting the homeless, who have recently been prioritised for the COVID-19 vaccination by the JCVI (39), and walk-in centres opening in community centres and places of worship, which could be replicated to engage groups such as undocumented migrants, refugees and asylum centres (40, 41). It is essential that vulnerable groups are made fully aware of such access points available to them, through collaborations with existing and trusted groups working in relevant communities, alongside scaling-up information campaigns to increase awareness of entitlement to register and use GP services (36). As well as supporting COVID-19 vaccine uptake, this could leave a lasting, positive impact on access to healthcare and confidence in vaccination going forwards, as marginalised communities are encouraged by the vaccination programme to come forward and register with primary care services.

Vaccine hesitancy issues were surprisingly common among participants, and mostly stemmed from a lack of accessible and understandable information, leading to concerns around vaccine contents, potential side effects and increased susceptibility to misinformation. An increased susceptibility to misinformation, often circulating on social media or by word-of-mouth, is known to be linked to an individual’s level of confusion, distress or mistrust around their social world (42). Our results suggest that a lack of accessible official information, social exclusion, and previous negative experiences with authorities (either health or political), may influence on susceptibility to misinformation. We have shown that hesitancy linked to circulating conspiracy theories was higher earlier in the pandemic (September – November 2020), before the start of the vaccination roll-out, suggesting recent messaging may have had some positive effect. These findings reflect similar findings from the Virus Watch study showing that 86% of adults in England and Wales (across all ethnic groups) who were reluctant or intending to refuse a COVID-19 vaccine in December 2020 had changed their mind in February 2021 (43). Hesitancy due to concerns around side-effects, vaccine contents and feeling clinical trials had been inadequate or too short, were voiced, particularly post-vaccine licensing (January-March 2021), with many expressing that they did not feel they had access to enough information. Another key influencing factor may be due to COVID-19 vaccine hesitancy and lower vaccine uptake in the early stages of the roll-out among healthcare staff from some ethnic minority groups (44, 45), who are often looked up to and trusted by their communities for health advice. Our results suggest that in precarious migrant groups, vaccine hesitancy issues could be relatively straightforward to address with clear, accessible and tailored information campaigns in a wide range of formats and languages. This should be done through existing schemes such as NHS community champions or Patient and Public Engagement groups (46) or through new collaborations with existing, trusted actors, such as charities, community groups and communities themselves, to ensure equitable uptake (14, 47).

Engaging precarious migrants, particularly undocumented individuals, in research has been rarely done to date, yet is essential to reveal unheard realities that these communities experience. Indeed, this study has shown that these groups may not be as ‘hard to reach’ as has historically been suggested, if appropriate communications channels are used (for example, through social media or trusted charities/community groups). However, our study has a number of limitations, including a lack of geographical representation from across the UK (most participants were resident in London or the North East). Whilst interpreters were available for participants, only two requested this service, meaning the study may have a bias towards those who have a higher level of English language. However, the interviews were designed to encourage discussion of participant’s wider community, meaning those with less language skills were often indirectly represented by their peers. Additionally, the researchers’ ethnicity and professional training may have influenced responses through perceived power differentials; the anonymous nature of telephone interviews, however, may have encouraged participants to share their views more freely.

This study has generated valuable insight into potential solutions and strategies to achieve equitable COVID-19 vaccine uptake among precarious migrants in the UK, with implications for other marginalised groups and with findings salient beyond the pandemic. More research is urgently needed to explore risk factors for low COVID-19 vaccine uptake in migrant and other vulnerable communities, to assess the extent to which barriers to access, vaccine hesitancy and circulating misinformation are playing a role. Research is now needed to ensure monitoring of equitable vaccine uptake in a wide variety of marginalised groups. Going forward, it will be critical that lessons learned during this pandemic around the importance of inclusiveness in health systems and principles of universal health coverage are embedded in the policy response, to improve access to health systems for excluded groups and to improve health outcomes in these growing populations in European and other high-income countries.

**Panel 1: Key messages and policy recommendations**

- Ensure strong and wide-reaching communication around strategies to support migrant populations to access COVID-19 vaccinations and the wider healthcare system – specifically undocumented migrants and those in high-risk settings such as asylum centres/accommodation – to ensure they are aware of options available to them and to allow equitable vaccine uptake in migrants currently outside of health systems.
- Implement accessible information campaigns in a wide range of formats and languages on COVID-19 vaccines (including side-effects, vaccine contents, counters to misinformation, entitlement and access points), delivered through trusted community sources (NGOs, community groups, religious groups, homeless centres, food banks). Information campaigns must be sensitive, culturally appropriate and must not risk stigmatising individual communities, which could negatively impact trust and engagement.
- Continue identifying new access points for the COVID-19 vaccine to ensure greater accessibility to the vaccine for precarious migrants and other excluded groups.
- Strengthen collaborations with local government, relevant charities and community groups, civil society groups, public health teams and healthcare professionals to develop engagement strategies with precarious migrant communities and other excluded groups to strengthen vaccine uptake. Actively involve communities in planning and implementation stages to develop trust and encourage widespread participation in COVID-19 vaccination programmes.
- Urgently conduct more research to explore risk factors for low COVID-19 vaccine uptake in migrant communities, to assess the extent to which vaccine hesitancy and circulating misinformation is playing a role, to better elucidate both individual and structural barriers to vaccination and strengthen monitoring to ensure equitable vaccine uptake in a wide variety of marginalised groups.
- There is a need to strengthen routine data systems in the UK and Europe to increase understanding around levels of access to health care, vaccination uptake, and health outcomes in diverse and growing migrant populations
- Ensure lessons learned during this pandemic around the importance of inclusion in health systems, through initiatives such as Universal Health Coverage, are meaningfully embedded in policy responses going forward

## Data Availability

The data will be made available on the St George's repository.

## Conflicts of interest

We declare that we have no conflicts of interest.

## Contributions

AD and SH had the idea for this study and designed the protocol. AD and SEH conducted interviews, AD, SEH, MN and OH carried out data analysis using a thematic framework developed by AD, with input from SH. AD, MN and SH contributed to a first draft of the paper. All authors viewed and discussed the data and contributed to the writing of the paper.

## Acknowledgements

We are grateful to the study participants who gave their time and shared their experiences and ideas, as well as members of our Project Board of migrant representatives for their valuable input. This work has been funded by the NIHR (NIHR300072). AD and SEH are funded by the MRC (MR/N013638/1). SH and AFC are funded by the NIHR (NIHR Advanced Fellowship NIHR300072) and the Academy of Medical Sciences (SBF005\1111). This work has been supported by the European Society of Clinical Microbiology and Infectious Diseases (ESCMID) Study Group for Infections in Travellers and Migrants (ESGITM). JC is funded by a National Institute for Health Research (NIHR) in-practice clinical fellowship (NIHR300290). FK is supported by a Health Education England / National Institute for Health Research (NIHR) Academic Clinical Fellowship. Kieran Rustage is funded by the Rosetrees Trust (M775). SMJ was funded by the National Institute for Health Research Health Protection Research Unit (NIHR HPRU) in Immunisation at the London School of Hygiene and Tropical Medicine (LSHTM) in partnership with Public Health England (PHE). AM is supported by the NIHR Applied Research Collaboration (ARC) NW London. The views expressed are those of the authors and not necessarily those of the NHS, the NIHR or the Department of Health and Social Care.

## Notes

### Competing Interest Statement

The authors have declared no competing interest.

### Author Declarations

The study was approved by the University of London ethics committee (REC 2020.00630).

